# The AbilityQuotient Dashboard: Outcomes of Implementing Patient-Specific Predictive Modeling in Inpatient Team Conference

**DOI:** 10.1101/2024.07.22.24310752

**Authors:** James A. Sliwa, Julia Carpenter, Andrew Bodine, Caitlin Deom, Richard L Lieber

## Abstract

**Objective:** Recent reports have highlighted the importance of data-driven decision making as it relates to precision medicine and the field of rehabilitation. One promising method to facilitate the integration of data into patient care involves the use of data warehousing to process and host patient data, analytics to produce useful results, and dashboarding technology to disseminate analytical results to care teams in a digestible and interpretable format. This report describes the implementation of a new composite rehabilitation outcome score (cROS), the AbilityQuotient, and predictive modeling into inpatient interdisciplinary conferences through a patient data dashboard and its impact on outcomes.

**Design:** 

**Setting:** Inpatient Rehabilitation Hospital

**Participants:** 13,397 patients completing inpatient rehabilitation from January 1, 2019 to December 31, 2023

**Intervention:** A patient centered, composite rehabilitation outcome score (cROS) and predictive modeling dashboard implemented into team conference

**Main Outcome Metrics:** Self-care and mobility IRF-PAI Form GG change scores, length of stay pre- and post-dashboard implementation; GG change scores compared to weighted national averages; clinician survey regarding perspectives of dashboard use; GG item long term goal modifications and goal attainment as a measure of influence on clinical plan of care

**Results:** After implementation of the patient outcomes dashboard into routine care, IRF-PAI Form GG self-care scores rose by 2.09 points and corresponding mobility scores rose by 7.18 points despite a 2.29 day reduction in length of stay. Further exploration investigating these changes as they pertained to payor revealed that these benefits occurred irrespective of insurer. Reports comparing facility to national averages extracted from eRehabData, a national outcomes data system and registry, suggest that the use of the outcomes dashboard resulted in greater reductions in length of stay and greater improvements in functional outcomes during the 2019-2023 period compared to the previous period. A corresponding survey assessing clinical perceptions of dashboard implementation revealed that it facilitated tracking and summarizing patient progress, reinforced the use of outcome metrics, and was perceived as valuable in goal setting and adjustment. Clinicians modified self-care goals six times more frequently and patients met these goals 19% more of the time after introduction of the cROS while they changed mobility goals nine times more frequently and patients met these goals 21% more of the time after introduction.

**Conclusion:** The incorporation of individual patient data and predictive modeling into rehabilitation patient care through use of a team conference dashboard has potential to provide an objective basis from which to perform precision rehabilitation. It also has the potential to impact outcome metrics improving value-based care and consequently deserves further study.

## Introduction

The ability to measure patient function and thus track progress during rehabilitation is required to demonstrate the benefit of interventions, establishing criteria for quality and value, and accurately making comparisons across patient groups, institutions, and post-acute care settings. There are established common data elements to measure status and change in self-care and mobility function in post-acute care, mandated as part of the Center for Medicare and Medicaid’s (CMS) payment and quality reporting programs. Additionally, there is emphasis from national professional organizations, private payors, institutions, and researchers to standardize outcome measure collection. Currently, such measures are used to demonstrate outcomes and value of rehabilitation services. Unfortunately, such data are rarely used to consider individualized prognoses, guide referral to post-acute levels of care, and monitor longitudinal progress (Butzer et al., 2019).

In contrast, the concept of precision rehabilitation is that the right treatment is applied at the right time to the right person, in the right setting (French et al. 2022). This philosophy emphasizes the importance of predictive modeling to enable clinicians to make data-driven treatment decisions based on an individual patient’s needs. Critical components of precision rehabilitation include the identification of patient subgroups who share specific characteristics related to disease progression and outcomes, standardized measurement of function, precise and frequent longitudinal measurement of real-world function, a comprehensive and well-maintained database, system and team science, and a focus on prediction rather than association (French et al., 2022). A consequence of using data to drive decision-making is that such data can be used for more than just reporting or insurance reimbursement but may help rehabilitation clinicians to actively alter their treatment plans and thus deliver more optimal outcomes for a specific patient.

Use of common data elements allow creates the capacity to harness electronic health record data to develop predictive models to support decision-making by clinical teams. While this is acknowledged to be an important step for supporting comparison of post-acute rehabilitation settings, and self-care and mobility data elements are required to be reported to CMS, they have measurement limitations that may limit their utility to achieve precision rehabilitation objectives. Specifically, these self-care and mobility functional items (and their predecessor, the FIM items) are ordinally scaled, which accounts for neither the multidimensional nature of functional tasks nor the differing degrees of difficulty of functional tasks (Merbitz, Morris, & Grip, 1989; Hobart, Cano, Zajicek, & Thompson, 2007; Liddel & Kruschke, 2018).

We previously addressed these limitations through development of AbilityQuotient (AQ) metric in the domains of self-care (Bodine et al., 2021), mobility (Deom et al., 2021), and cognitive-communication (Carpenter et al., 2024) that are computed with functional item level data from mobility/self-care IRF-PAI section GG (Centers for Medicare & Medicaid Services, 2015) or cognitive Functional Independence Measure (Granger, Hamilton, Keith, et al., 1986), as well as additional standardized assessment data (all of these items are found in Supplemental Tables 1-3). Inclusion of the additional standardized assessment data in the AQ allows for more precise measurement of patient status and change in function, with improved sensitivity to detect change and improved measurement for more and less severely impaired patients. This published work described selection and standardization of assessment data from over 8,000 rehabilitation patients. Confirmatory factor analysis (CFA) yielded models for each domain, thereby delineating assessment items that comprise self-care, mobility, and cognitive-communication domains. Then, multidimensional item response theory (MIRT) methods was used to create overall scores for each domain, as well as subscores for subcomponents of function. For example, the AQ Self-Care assessment’s measurement model can yield scores for overall self-care in addition to balance, upper extremity function, and swallowing. These MIRT-derived scores comprising the AbilityQuotient (AQ) have means of zero with scores spanning negative (below average patient ability) and positive (above average) values which represent a continuous range of patient function.

As an illustration of the shortcomings of ordinal scales used in QI and FIM, AQ scores can be compared to these scales by computing the cut-points between ordinal scale rating categories over the continuous range of ability. For example, as seen in Figure 1A, an AQ score of 0.0 would likely correspond to a QI rating of 3 (partial/moderate assistance) on chair to bed transfers. Yet, the AQ demonstrates superior psychometric properties in comparison to the FIM and QI scales as noted by its greater sensitivity compared to traditional ordinal categories (Bodine et al. 2021, Deom et al. 2021). For example, as seen in Figure 1B, a patient’s chair/bed transfer score does not change intervals on the QI’s rating scale, while it does change from a -2.0 to -1.0 on the AQ. The AQ also has higher ceilings and lower floors compared to the FIM or QI, which better captures change for less and more severely impaired patients. Additionally, the AQ’s MIRT scores reflect a continuous scale of ability, which normalizes how change is captured. For example, the amount of improvement in mobility made by a patient moving from a –2.0 to –1.0 or –1.0 to 0.0 on the AQ’s metric is one point irrespective of initial severity; in contrast, the amount of improvement made on the chair/bed transfer item’s ordinal scale depends heavily on initial severity (Fig. 1C). A more impaired patient progressing from –2.0 to –1.0 on the AQ would not improve on this item, but a comparatively less impaired patient progressing from –1.0 to 0.0 could expect to proceed from category 1 (dependent) to category 3 (partial/moderate assistance). A similar region of insensitivity can similarly be seen for patients towards the higher end of the metric as well. In short, scoring over a standardized, continuous range of ability permits more precise and sensitive assessment compared to current measures.

**Figure 1.**
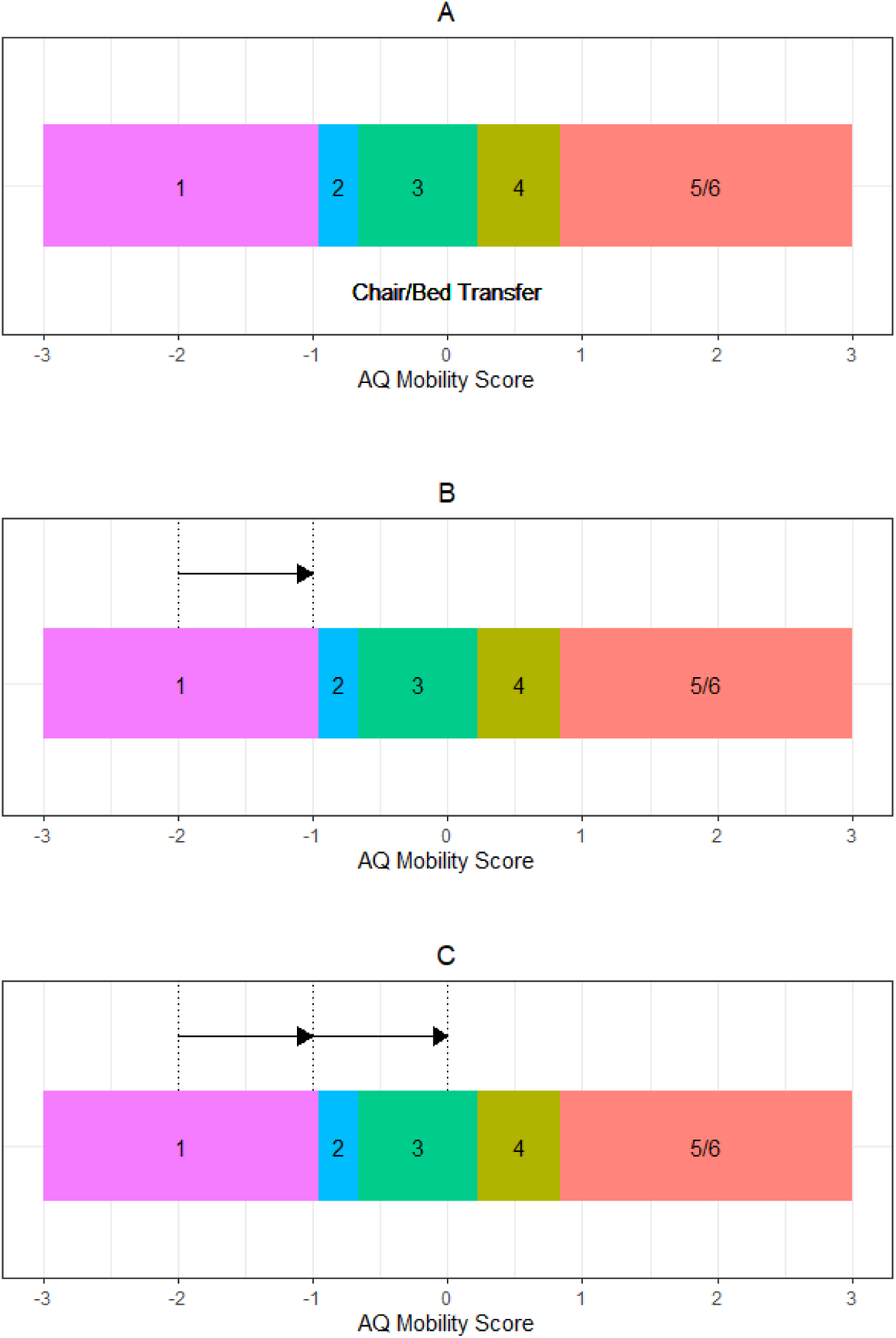
A): Comparison of functional ratings on the ordinally-scaled IRF-PAI Form GG item “Chair to Bed Transfer” to the continuous MIRT scoring scale for the mobility domain. B): Demonstration of MIRT score change not being captured by functional scale. C): Demonstration of equally-sized increases on continuous MIRT scale suggesting different amounts of functional gain on the ordinal rating scale

While an AQ score in a given domain demonstrates a patient’s composite score over time, such a score can also be used to develop predictive models for groups of patients. Group-based predictions can forecast probable performance over an episode of care for diagnostically-defined sets of patients, and comparison of such a group-based prediction to a patient-specific individual curve can determine whether an individual patient will or will not meet projected levels by a given discharge date. This type of information allows the clinical team to link data to an individual’s clinical course, review predictive measures for a given patient, and monitor the course of recovery relative to prognosis in real time. However, for outcomes data and predictive modeling to drive decision-making, clinicians and interdisciplinary teams must be able to quickly visualize actionable data.

Consistent with the need to make rapid decisions based on high quality data, is the growing use of dashboards in healthcare. Health care dashboards are typically employed to serve operational, quality, safety, or practice improvement initiatives (Bucalon et al., 2022). They make use of structural, progress, or outcomes related data at either generic or disease-specific levels. A dashboard for interdisciplinary conference that uses patient-specific predictions has the potential to help teams make tailored decisions.

We sought to align with precision rehabilitation objectives through the deployment of a dashboard that visualizes scoring and predictive modeling results in inpatient interdisciplinary team conference. More specifically, our aims were to 1) design a dashboard with clinical stakeholders that incorporate the AQ metric and predictive models, 2) implement and assess the impact of the dashboard on clinician perceptions and plan of care, and 3) evaluate the effect of dashboard implementation on patient outcomes and length of stay as markers of value-based care.

## Methods

### Design of AbilityQuotient (AQ) Dashboard

The purpose of using a dashboard to display the AQ metric and predictive models was to provide the interdisciplinary rehabilitation team with current data on patient progress and the ability to compare score changes over time to a predictive model, fostering team discussion and plan of care decision-making. Design of the dashboard was achieved by engaging a multi-stakeholder group, a beta-version pilot used to gather clinical feedback, and integration of that clinical feedback into the final design. Initially, a team composed of an inpatient rehabilitation physician, a clinical manager with a physical therapy background, the hospital’s chief medical officer, and a psychometrician met to discuss design and deployment of a dashboard for clinical use. A prototype version was created using the Shiny package (version 1.0.4) for the R programming language (Chang et al., 2017). It included specific features in (** in Table 1) and was introduced to an interdisciplinary team consisting of physical and occupational therapists, speech-language pathologists, nurses, and care managers to trial for team conference with the aforementioned physician. After an introductory session to review the dashboard and models, discipline-specific breakout groups met with trainers to discuss clinical cases and pilot team conference communication with the use of the prototype. This pilot was informative to identify clinical training and team conference communication needs, suggest modifications to graphics, and reduce the complexity of the data displayed. After the pilot study dashboard introduction, teams noted shortcomings of a diagnosis-focused predictive model since patients admitted at a higher or lower level of ability than average for that diagnosis quickly outpaced or demonstrated limited ability to meet the diagnostic prediction. The predictive model was re-envisioned as a set of nested piecewise hierarchical linear mixed models (HLMMs) with either random slopes and piecewise intercepts to provide patient-specific predictions or only random slopes to provide an adjusted diagnosis prediction. Features were added to the dashboard following the pilot (++ in Table 1).

**Table 1.**
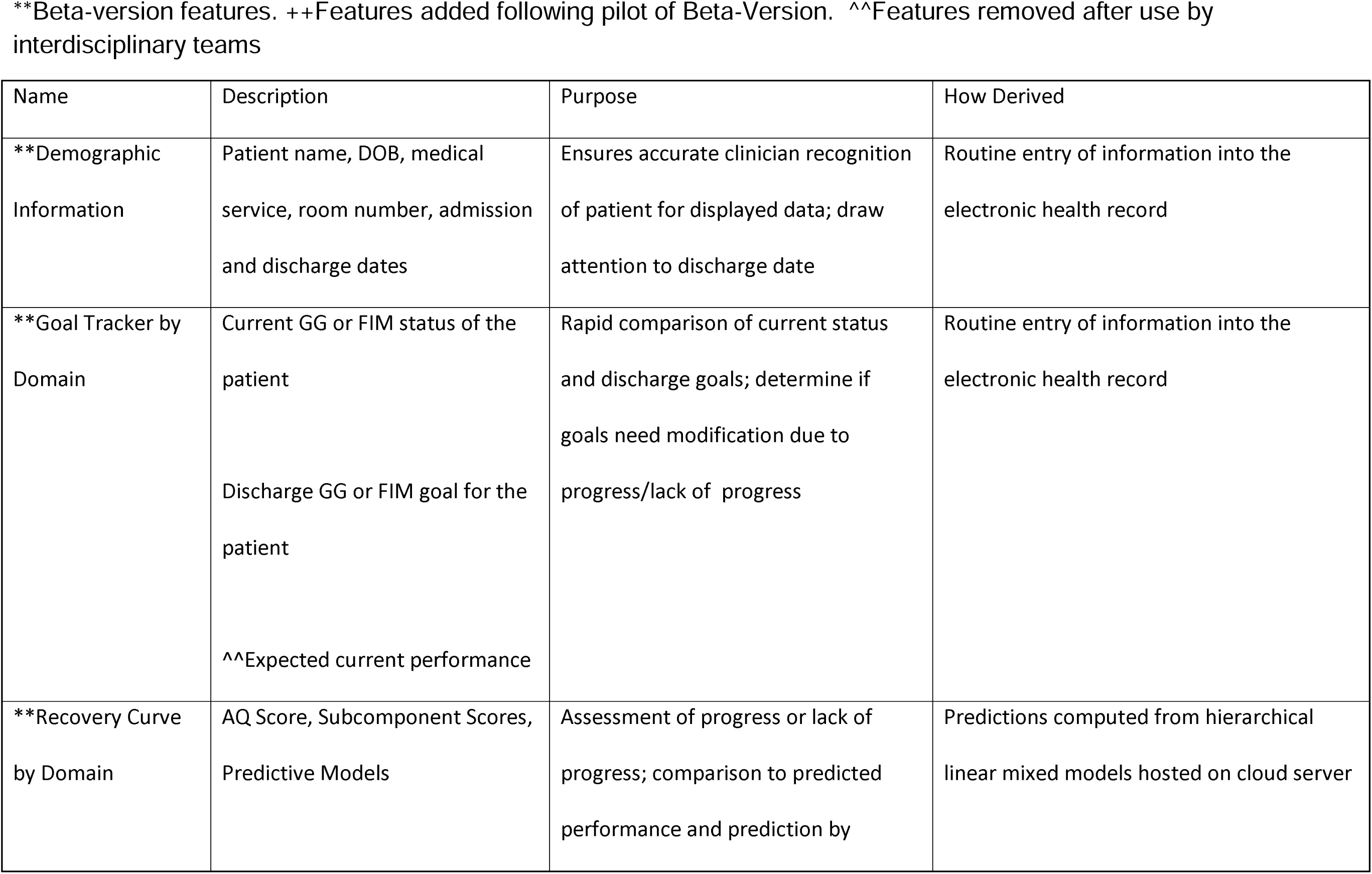
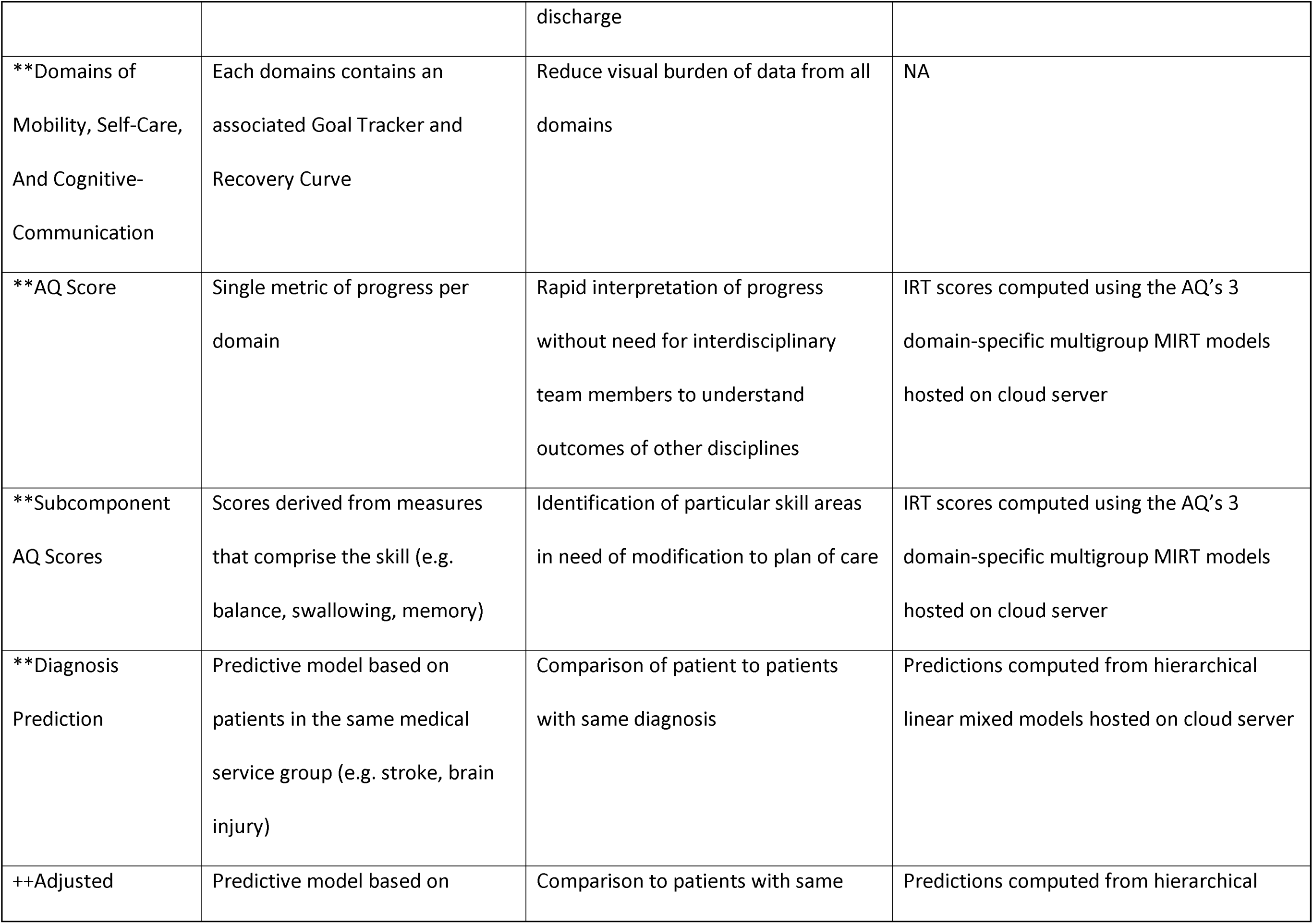
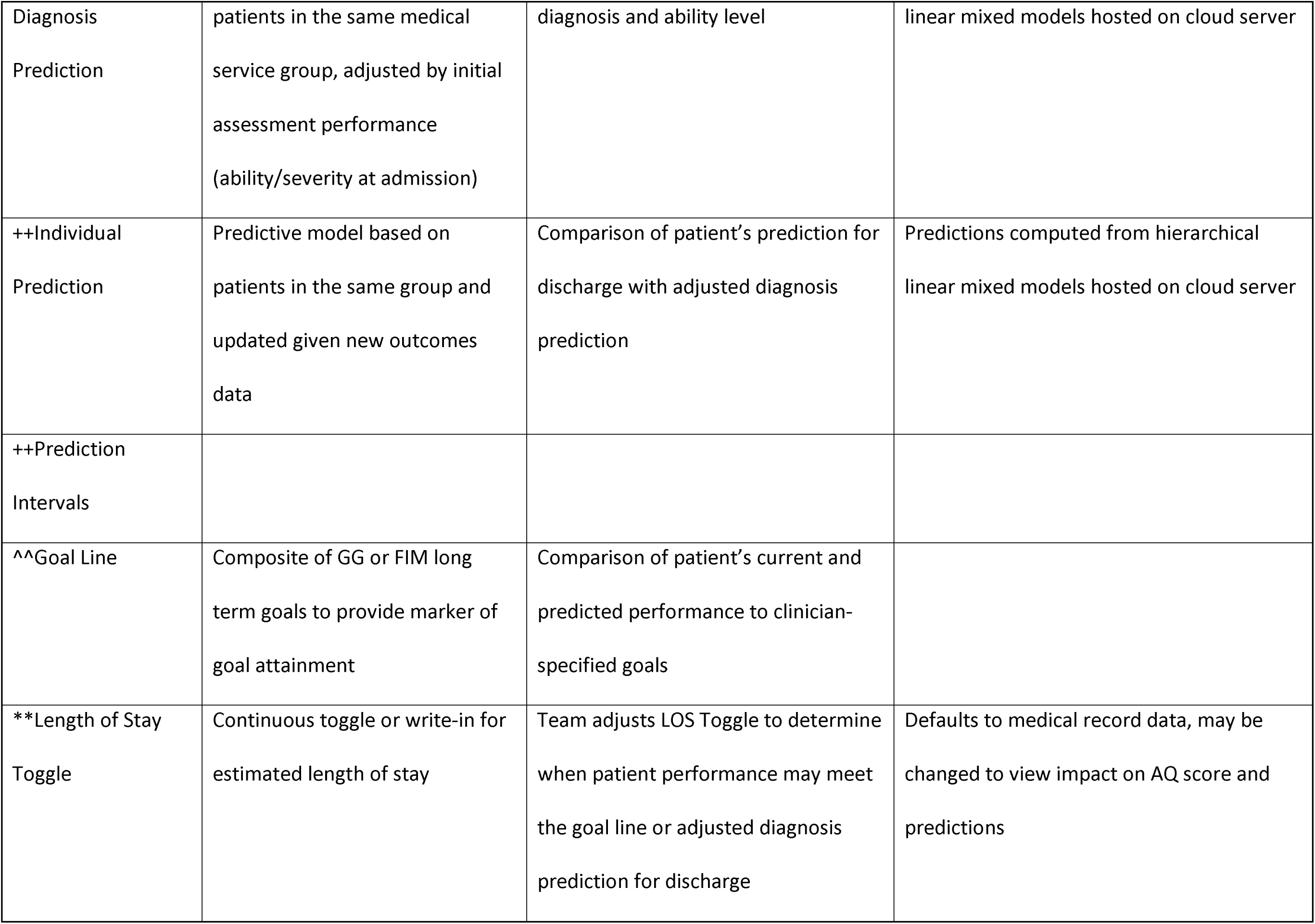
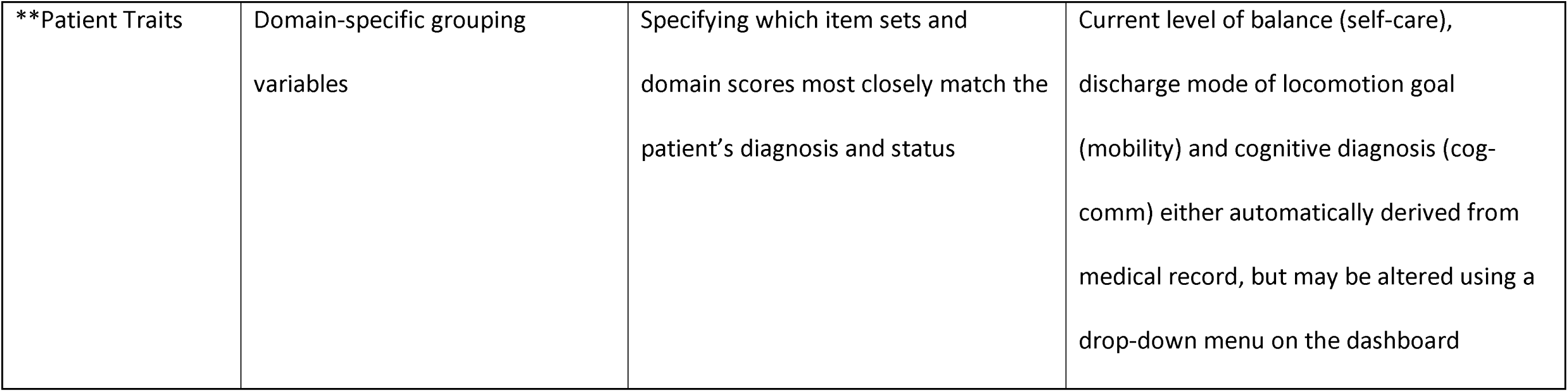
Ability Quotient Dashboard Features.

Next, the development of the dashboard included additional rehabilitation team members, leadership from the hospital Information Systems team, clinical directors, and consulting firms with expertise in database administration, cloud systems engineering, and web development. A key requirement for the AQ Dashboard was rapid integration of novel patient assessment data, and this need was met through the development of automated data pipelines that transmitted data from an electronic health record (EHR), through an enterprise data warehouse (EDW), and finally into a high-availability application database on a daily basis. Once new data were loaded into the application database, R programming language scripts were triggered to compute scores and either generate or update predictions for the daily data import.

In addition to developing data systems and implementing design characteristics, this group tested the logic of the AQ Dashboard using real patient data in a test environment to identify and rectify issues. The next iterations of the AQ Dashboard, specifically Goal Tracker (Fig. 2) and Recovery Curve Sections (Fig. 3) were developed. Importantly, because the AQ is composed of multiple clinical assessments, the underlying basis for any change in the overall score can be “drilled down” with fairly high granularity (Fig. 4). Certain features. were ultimately removed from the Dashboard after a larger rollout and feedback from interdisciplinary teams (labelled ^^ in Table 1). The Goal Line was ultimately removed because it failed to provide a meaningful comparison point to the patient’s AQ score. This was because the MIRT score associated with the Goal Line was derived solely from QI items, rather than all items comprising the AQ score. The net result of the very large analytical effort on our patient data was to create predictive models that were diagnosis and severity specific so that any patient’s individual AQ scores (Fig. 5, black line) could be compared to a diagnostically similar group (Fig. 1, gray line).

**Figure 2.**
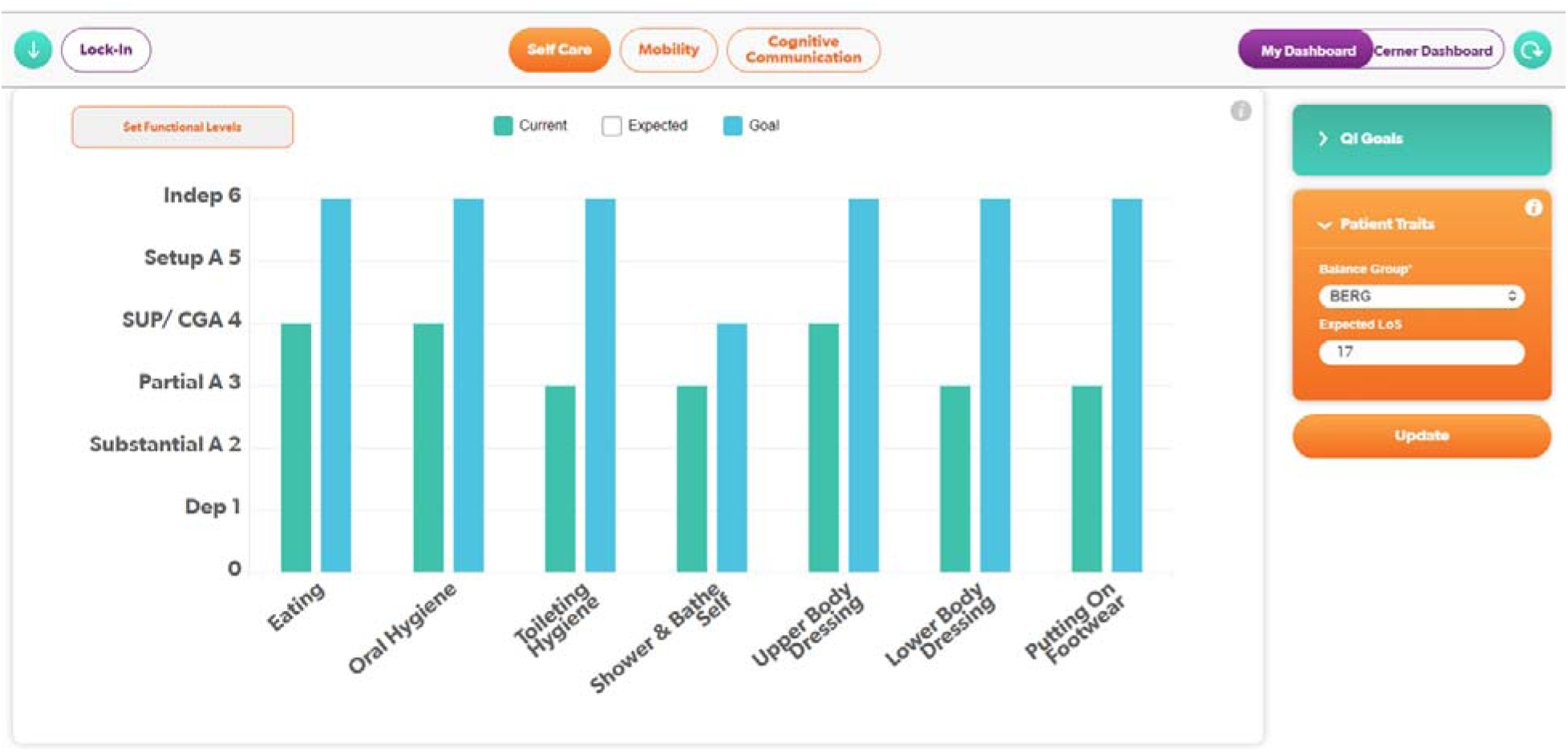
Goal Tracker section of the AQ Dashboard displaying current status and long term goals for GG or FIM items.

**Figure 3.**
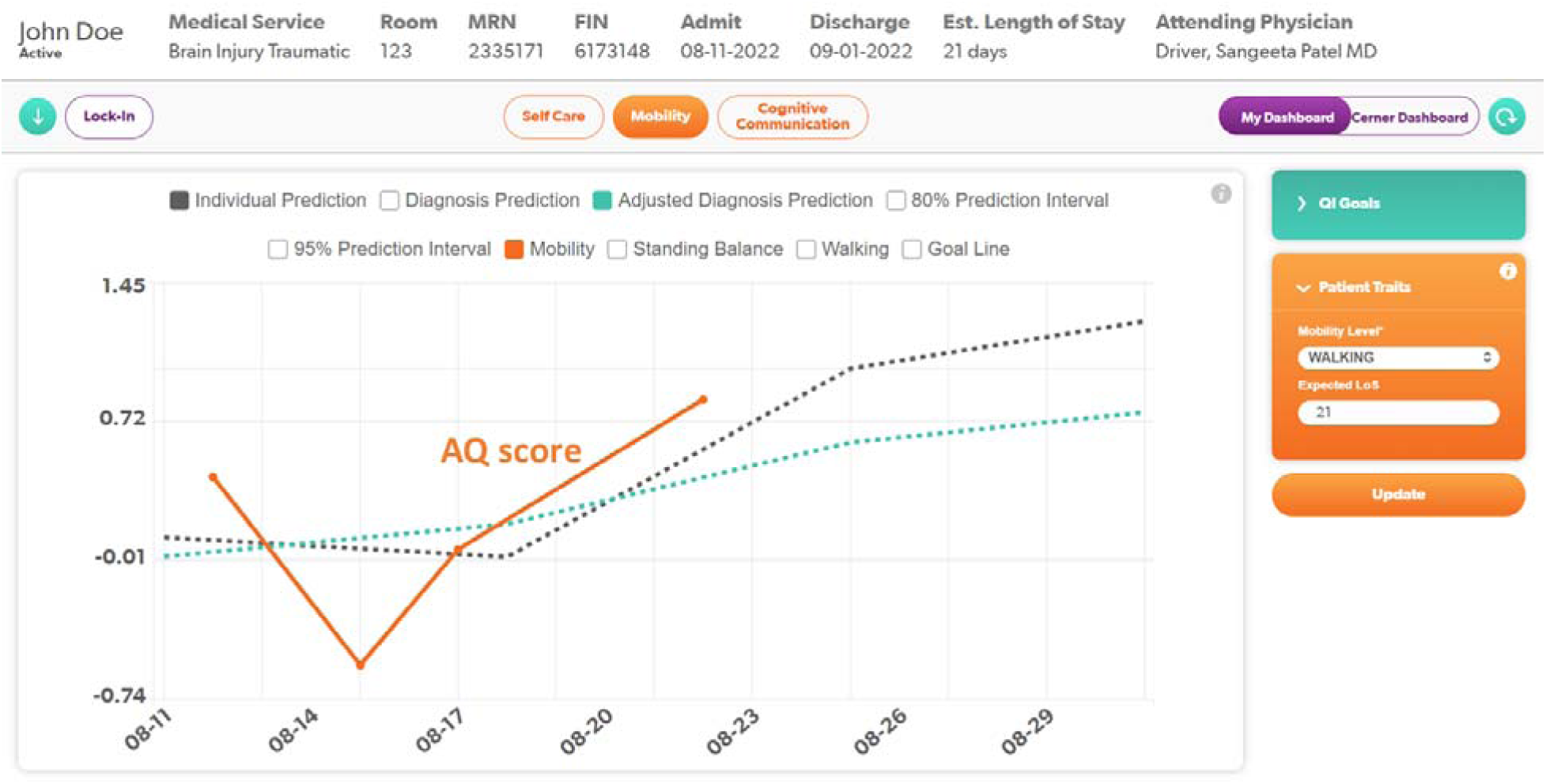
Recovery Curve section of the AQ Dashboard displaying AQ score, adjusted diagnosis prediction, and individual prediction.

**Figure 4.**
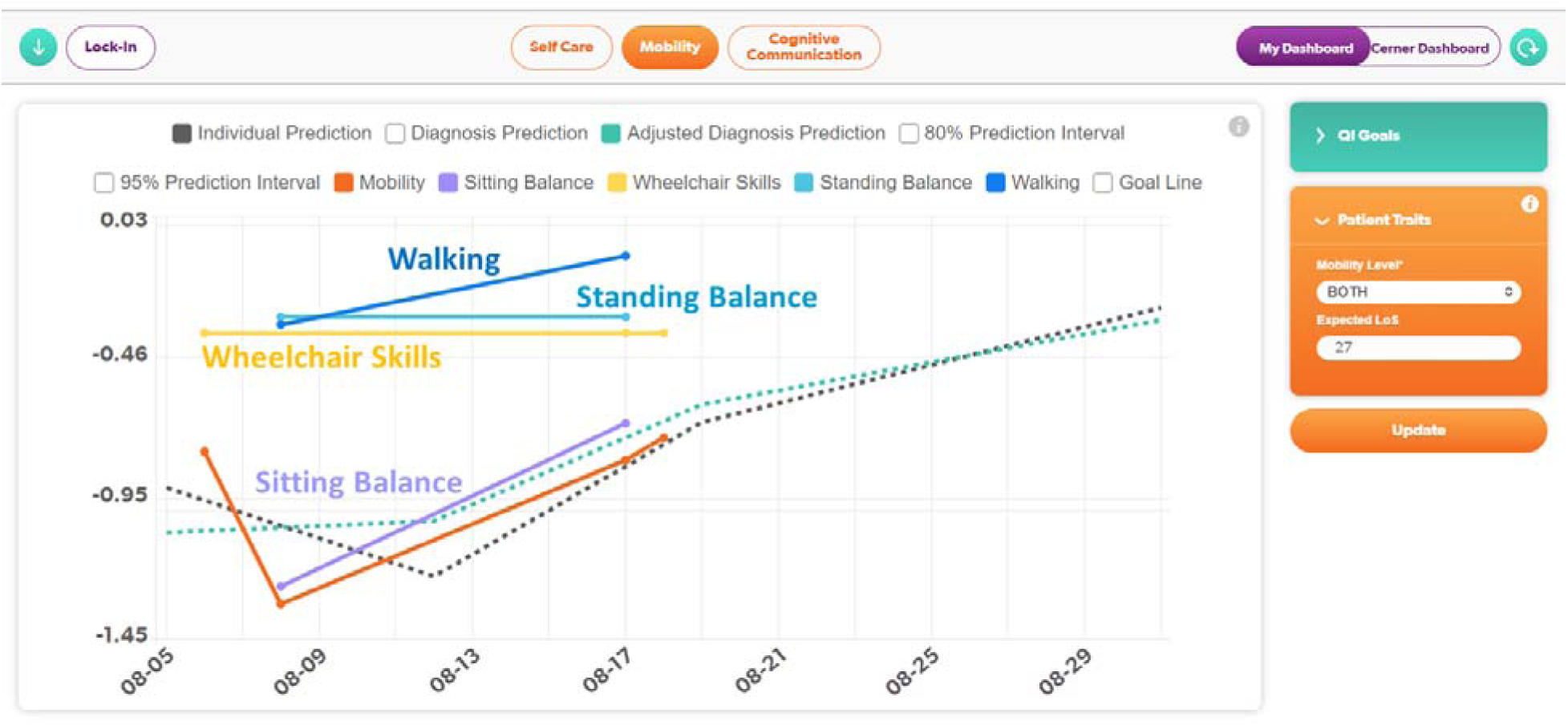
Recovery Curve section of the AQ Dashboard displaying subcomponents of function in the Mobility Domain.

**Figure 5.**
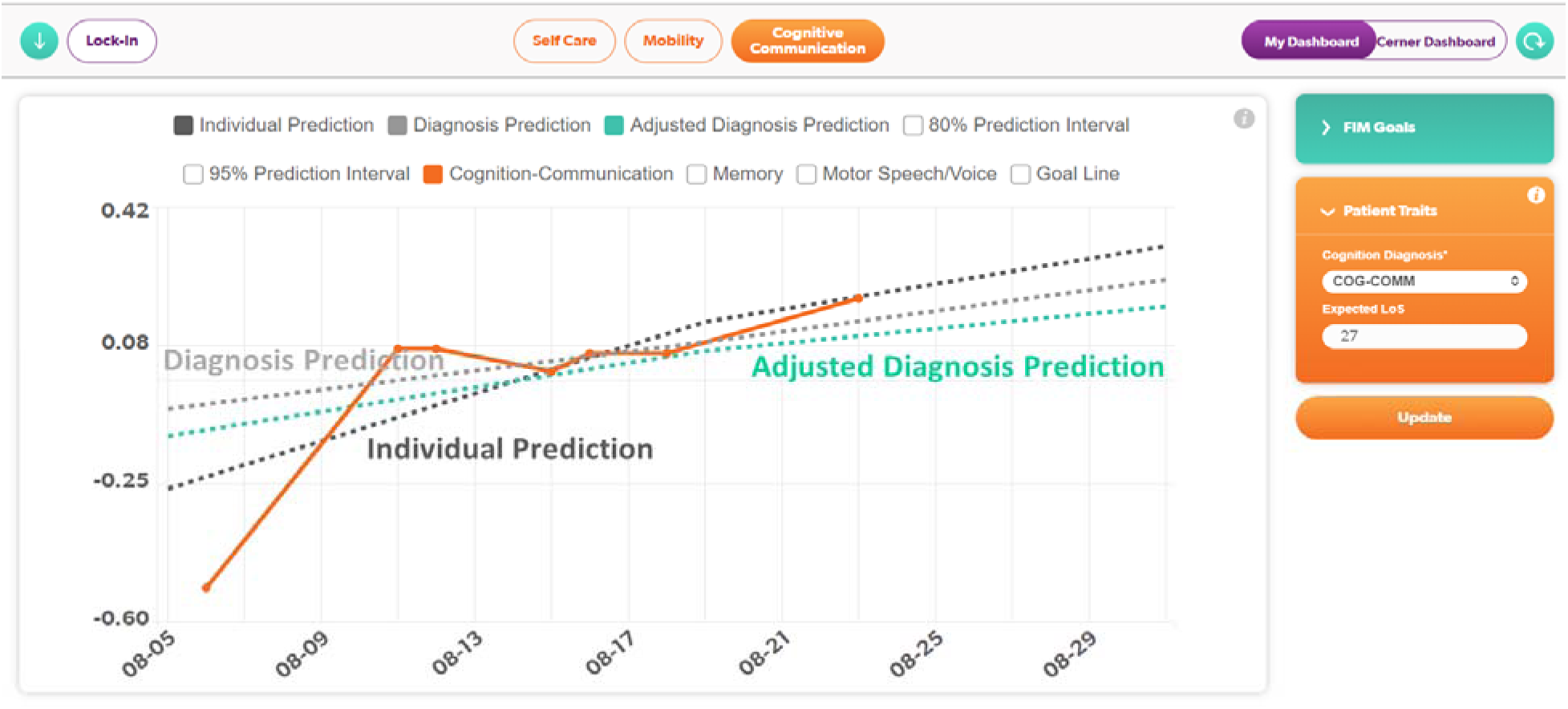
Recovery Curve section of the AQ Dashboard displaying all predictive models (diagnosis prediction, adjusted diagnosis prediction, and individual prediction)

### Clinical Implementation and Assessment of Clinician Perspective

Implementation of the AQ dashboard into team conference required a shift in interdisciplinary communication and workflow. Typical inpatient conferences prior to use of the AQ Dashboard involved discussion patient current status and goals in GG or FIM scales, identification of factors limiting progress, interdisciplinary management needs, discharge planning. Using the AQ Dashboard in team conference required interpretation of data and incorporation of that data into team-based treatment decisions and planning. Consequently, it necessitated an organized and structured initiative to train physicians, nurses, and allied health staff.

Our inpatient rehabilitation hospital has seven inpatient floors (three treating stroke, brain injury, other neurologic diagnoses; two treating spinal cord injury; and two treating general rehabilitation diagnoses) for a total of 262 beds. From June 2020 to November 2020, education was implemented sequentially by unit and began with unit leadership including attending physicians and therapy managers. Standardized training included clinical interpretation of the AQ dashboard, shaping team conference communication using the dashboard, and implementing workflow and practice changes. Based on feedback from the pilot, scaffolded training sessions and objectives were conducted for physical therapists, occupational therapists, and speech-language pathologists. They participated in an 8-session series that involved practice and feedback; The training schedule and progressive session learning objectives were structured (Table 2). Sessions were either discipline-specific or team-based, with integration of the AQ Dashboard into live team conferences halfway through the training. To ensure translation of practiced knowledge and skills into team conference, trainers completed regular audits, which fostered specific, real-time feedback for managers and clinicians. Finally, to sustain these training efforts within an evolving workforce, on-demand training content was created to support staff learning about the AQ dashboard and how to incorporate its data into team conference communication. Additionally, Innovation Center (inpatient) allied health clinical educators provide ongoing support for interpretation and guidance for team conference.

**Table 2.** Training Sessions for Physical Therapists, Occupational Therapists, and Speech-Language Pathologists.

| Session | Duration/Type | Objectives |
| --- | --- | --- |
| #1 Introduction | 18 minutes<br>On-Demand E-Learning | 1. Discuss clinical rationale for development of the AQ metric and Dashboard<br>2. Define basic components of the AQ Dashboard<br>3. Describe intended uses of the AQ Dashboard<br>4. Outline training plan for allied health clinicians |
| #2 Discipline Specific Overview | 45 minutes<br>Live | 1. List the measures that contribute to the AQ for their discipline<br>2. Identify two primary components of the AQ Dashboard: Goal Tracker and Recovery Curve<br>3. Identify parameters during modeling of AQ team conference<br>4. Explore AQ Dashboard domains, Goal Tracker, and Recovery Curve |
| #3 Discipline Specific Practice Cases | 1 hour, 45 minutes<br>Live | 1. Describe impact of Patient Traits on the AQ<br>2. State appropriate uses of “If-then” vs. Lock-In features on AQ Dashboard<br>3. Following a model, interpret the Goal Tracker and Recovery Curve for a given patient for initial and interim conferences<br>4. Evaluate timing of team conference communication |
| **Team Begins Use of AQ Dashboard in Team Conference** |  |  |
| #4 Discipline Specific Cases from Current Caseload | 1 hour, 45 minutes<br>Live | 1. Verbalize non-AQ patient considerations for team conference<br>2. Appropriately review the Goal Tracker, Recovery Curve, and non-AQ considerations for a given patient (initial and interim as available)<br>3. Write AQ team conference coverage for a sample patient<br>4. Discuss decision-making considerations given data presented on Dashboard |
| #5 Team Practice Cases | 45 minutes<br>Live | 1. Interpret the Goal Tracker for your team (initial and interim)<br>2. Interpret the Recovery Curve by domain for team audience (initial and interim)<br>3. Address non-AQ patient considerations as a team |
| #6 Discipline Specific Cases from Caseload | 45 minutes<br>Live | 1. Describe impact of QI/FIM and assessment scores on Recovery Curve trends<br>2. Efficiently interpret the Goal Tracker, Recovery Curve, address non-AQ patient considerations |
| #7 Team Feedback | 45 minutes<br>Live | 1. Self-assess/compare current performance with targeted performance for reporting during AQ conference |
| #8 Discipline Specific Question and Answer | 45 minutes<br>Live | 1. Resolve outstanding questions<br>2. Evaluate own adherence to team conference parameters and additional learning needs |

After this initial training phrase, teams participated in two additional sessions in March and April 2021 that used a decision-tree to support interpreting the AQ dashboard for clinical decision-making related to the plan of care (Fig. 6). Finally, after audit of team conferences SCI and NMB teams were identified for additional sessions, specifically focused on physician involvement in team conference and interdisciplinary communication to balance interpretation of the dashboard as well as discharge planning needs.

**Figure 6.**
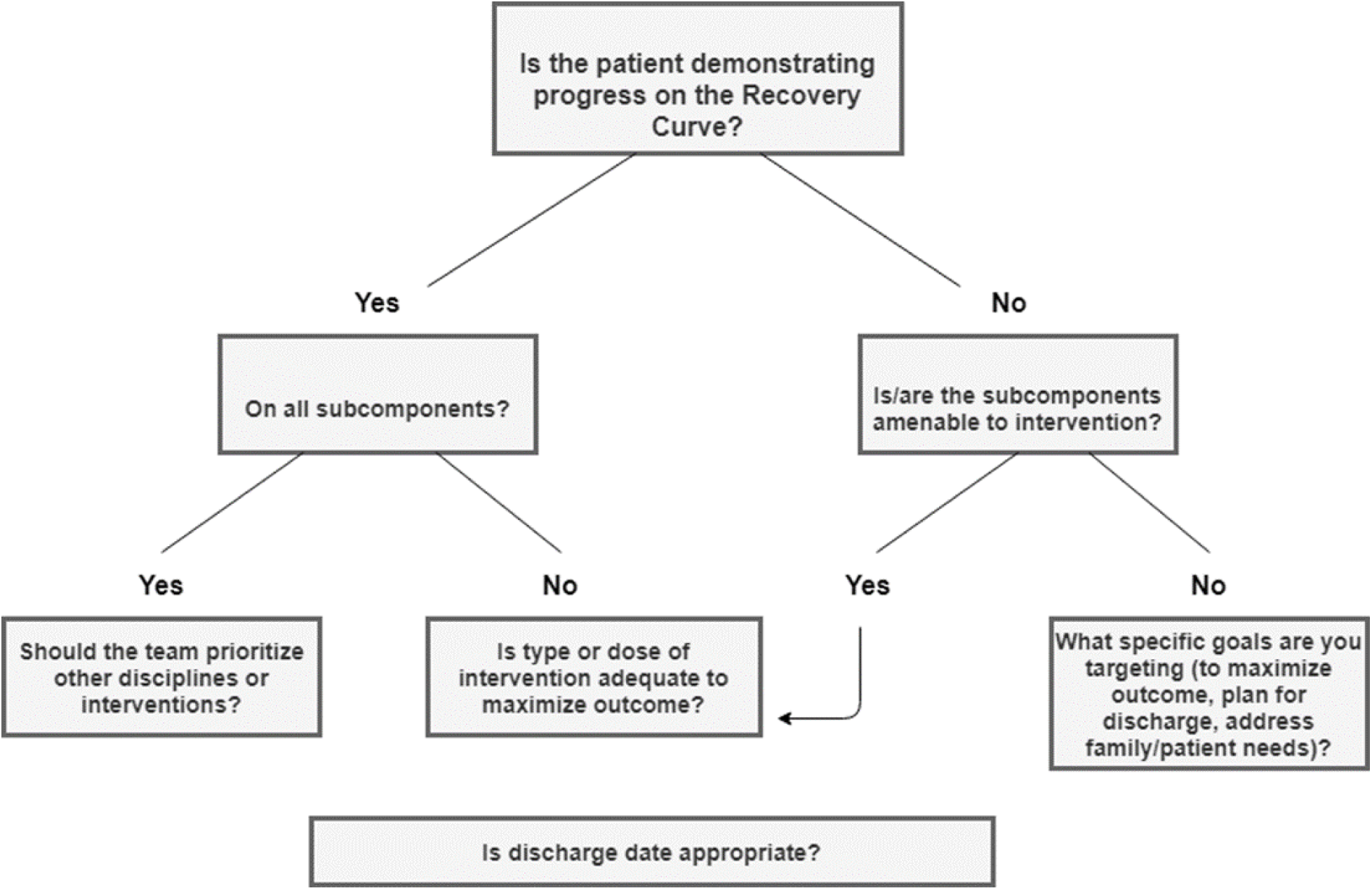
Decision-tree for decision-making with use of the AQ Dashboard.

Clinician perspectives on use of the AQ Dashboard were gathered via survey to the interdisciplinary teams. At 21 months post-implementation, a survey of all disciplines was conducted to evaluate acceptance and assess the AQ Dashboard’s impact on clinical practice. Survey questions were divided into six categories, including the perceived value of incorporating the AQ into team conference, its influence on team communication and plan of care, overall value, feasibility of integrating assessment tools into practice, and an overall summative evaluation. Survey questions were rated on a 5-point Likert scale using the ratings Strongly Agree, Agree, Neutral, Disagree, and Strongly Disagree.

Survey creation, administration, and data collection were performed using Redcap (Harris et al., 2009; Harris et al., 2019). Clinicians were permitted one month to respond and sent reminders, after which the data were extracted from Redcap and prepared for analysis using the R programming language. In total, 115 responses (35 PTs, 23 OTs, 13 SLPs, 12 care managers, 5 therapy managers, 15 physicians, and 14 nurses) were recorded for the 18-item survey. Survey rating scale data were converted to an integer value of -2 (Strongly Disagree) to 2 (Strongly Agree) for the sake of computing comparable average item ratings such that negative values suggest negative affect, positive values reflect positive affect, and averages near 0 imply neutrality or indifference.

Additionally, to understand how clinicians changed their plan of care after AQ Dashboard implementation, we measured the frequency at which clinician’s changed their GG item level long term goals during the clinical encounter as well as attainment of those goals pre- and post-AQ dashboard implementation. *Assessment of Patient Outcomes and Value Based Care*

To determine the impact of dashboard implementation on patient outcome measure performance, changes in IRF-PAI Form GG self-care and mobility subscores were extracted from the electronic health record. Form GG performance was used as the comparative measure due to 1) therapists having access to Form GG scores for their patients prior to the dashboard’s roll-out, 2) greater recognition of the IRF-PAI within the field of physical rehabilitation, and 3) the availability of national Form GG averages from industry registries and outcomes systems like eRehabData. “Transitionary” patients present at the hospital during the June 2020-November 2020 clinician training period were excluded from the sample if their episode of care overlapped with their clinician’s training phase.

To reduce the impact of extraneous factors on the analytical results, age, biological sex, length of stay, and admission Form GG scores were used as covariates in a series of regression analyses investigating year-over-year changes in Form GG performance improvement using estimated marginal means (EMMs) with the Bonferroni adjustment (Draper and Smith, 1981). These results used length of stay, Form GG self-care change, and Form GG mobility change as dependent variables. EMM comparisons for the provider-specific results were further computed over the margin of provider.

In addition to the results described above, yearly reports detailing facility and national Form GG averages from the years 2019 through 2023 were extracted from eRehabData (eRehabData, 2024). Although these tables do not contain mean comparisons due to the lack of standard deviation or standard errors within the reports, the pattern of means comparing Form GG gains over time could still be used to identify instances in which clinical dashboard usage appeared to offer advantages compared to other facilities.

## Results

### Clinician Perspectives and Long Term Goals

Most positive responses were seen for several surveyed areas as defined by an average item rating greater than .30 (Table 3). These included a positive impressions of the AQ Dashboard regarding its influence on setting or adjusting goals (.49), completion of measures in order to track patient progress (.43), tracking the progress at the domain level to summarize progress (.41), ease of interpretation (.36), reinforcing outcome measure use (.35), and use of the predictive models to help understand if a patient’s progress is on track (.30). Clinicians also had a positive impression regarding whether the measure that comprise the patient’s AQ score were feasible to complete during their rehabilitation stay (.46).

**Table 3.** Average item rating of survey responses after conversion of Likert Scale to –2 to 2.

| Item | Overall |
| --- | --- |
| Setting or adjusting goals | 0.49 |
| Completing the repeated measures that comprise the patient's AQ score is feasible to do throughout the patient's IP rehabilitation stay | 0.46 |
| Completing the AQ outcome measures is important to track patient progress | 0.43 |
| Tracking progress at the domain level (Self-care, Mobility, Cognitive-Communication) facilitates summarizing progress | 0.41 |
| The visual format and metrics in the AQ Dashboard are easy to interpret | 0.36 |
| Knowing that data will be discussed in team conference reinforces my use of outcome measures in routine practice | 0.35 |
| Using predictive models helps me understand if my patient's progress is on target | 0.30 |
| Makes team conference outcomes focused | 0.17 |
| Facilitates my thinking about setting patient goals and patient progress | 0.15 |
| Informing discharge date | 0.13 |
| Completing the AQ outcome measures is important to inform my treatment plan | 0.13 |
| Making Clinical decisions | -0.03 |
| Highlights key information for discussion during team conference | -0.07 |
| Facilitates a better outcome for patients | -0.13 |
| Use of the AQ Dashboard enhances the efficiency of team conference | -0.17 |
| Enhances goal achievement because it allows the team to prioritize and change the dosing of therapeutic interventions | -0.21 |
| When compared to team conference prior to its use, the AQ has improved team conference | -0.29 |
| Identifying and addressing barriers to discharge | -0.43 |

Most negative responses as defined by negative average values were seen for items regarding the AQ Dashboard’s influence on making clinical decisions (-.03), its ability to highlight key information during conference (-.07), its ability to facilitate a better outcome for patients (-.13), impact on conference efficiency (-.17), influence on goal achievement (-.21), improvement compared to previous team conference format (-.029), and ability to identify and address barriers to discharge (-.43).

Neutral responses were provided as defined by averages between .3 and 0.0 for items regarding the AQ Dashboards focusing of conference on outcomes (.17), facilitating thinking about goals and progress (.15), informing the discharge date (.13), and whether completing outcomes is important to the treatment plan (.13).

Clinicians changed their long term goals for GG items more frequently in the post-implementation sample. They modified their goals six- and nine-times more frequently in self-care and mobility, respectively. Goal attainment rates after AQ implementation for GG item long term goals in the self-care and mobility domains also increased by 19 and 21%, respectively.

### Clinical Outcomes and Value Based Care

Over the course of the five-year window, length of stay decreased by an average of 2.29 days, Form GG self-care subscores improved by 1.98 points, and Form GG mobility scores improved by 7.18 points (Table 4). While the pattern of significance does not indicate consistent year-over-year improvements, all three outcome metrics suggest more favorable results for the most recent year compared to 2019.

**Table 4.** Year-by-year length of stay and change scores for self-care and mobility domains.

| Year | N | LoS | ΔSelf-Care | ΔMobility |
| --- | --- | --- | --- | --- |
| 2019 | 2,500 | 21.27 | 12.67 | 26.96 |
| 2020 | 2,566 | 19.91* | 14.03* | 32.37* |
| 2021 | 2,739 | 19.54* | 14.26* | 32.86* |
| 2022 | 2,732 | 19.79* | 14.55* | 32.94* |
| 2023 | 2,860 | 18.98* | 14.76* | 34.14* |

\* = Significantly different from 2019 value

We further investigated whether significant changes across length of stay, self-care functional scores, and mobility functional scores were present for patients insured by either Medicare or commercial providers (Table 5). The results suggest that both types of payor saw significant decreases in length of stay (Medicare: -1.98 days; Commercial: -2.44 days) and significant increases in IRF-PAI form GG scores (Medicare, self-care: +1.98 points; Medicare, mobility: +6.67 points; Commercial, self-care: +2.11 points; Commercial, mobility: +7.01 points) between 2019 and 2023.

**Table 5:** Changes in length of stay, IRF-PAI Form GG Self-Care, and IRF-PAI Form GG Mobility during the study period, separated by provider (Medicare or commercial)

| Year | Provider | N | LoS | ΔSelf-Care | ΔMobility |
| --- | --- | --- | --- | --- | --- |
| 2019 | Medicare | 1,293 | 20.05 | 12.22 | 26.15 |
| 2020 | Medicare | 1,198 | 19.33 | 13.51* | 31.19* |
| 2021 | Medicare | 1,260 | 18.67* | 13.67* | 31.59* |
| 2022 | Medicare | 1,189 | 18.79* | 13.97* | 31.81* |
| 2023 | Medicare | 1,261 | 18.07* | 14.20* | 32.91* |
| 2019 | Commercial | 1,831 | 21.49 | 12.64 | 27.19 |
| 2020 | Commercial | 1,943 | 19.90* | 14.21* | 32.90* |
| 2021 | Commercial | 2,038 | 19.64* | 14.38* | 33.39* |
| 2022 | Commercial | 2,075 | 19.70* | 14.65* | 33.35* |
| 2023 | Commercial | 2,038 | 19.05* | 14.75* | 34.20* |

The eRehabData-derived metrics for the local facility were compared to national weighted averages (Table 6). Because these results were pulled directly from an eRehabData report and were not adjusted using the same covariates as the facility-specific year-over-year results described above, there is no expectation that the results in Table 5 would match those in Table 4. The “overall” results suggest that between 2019 and 2023, the facility using the clinical dashboard saw a greater reduction in length of stay (by 1.77 days) while enjoying larger gains in self-care (by 0.78 points) and mobility (by 3.19 points).

**Table 6.** Facility vs. Weighted National Average (eRehabData) length of stay and change scores for self-care and mobility domains.

| Year | Facility |  |  | Weighted National Average |  |  |
| --- | --- | --- | --- | --- | --- | --- |
| | LoS | ΔSelf-Care | ΔMobility | LoS | ΔSelf-Care | ΔMobility |
| 2019 | 22.38 | 10.10 | 21.15 | 15.48 | 9.80 | 23.08 |
| 2020 | 22.46 | 11.17 | 24.82 | 16.10 | 10.52 | 23.93 |
| 2021 | 21.80 | 11.68 | 25.63 | 15.63 | 10.83 | 24.65 |
| 2022 | 21.28 | 11.80 | 25.85 | 15.27 | 10.79 | 25.10 |
| 2023 | 20.61 | 12.03 | 26.99 | 14.87 | 10.95 | 25.73 |

## Discussion

In this quality improvement project, we implemented a patient-specific predictive model and outcomes dashboard in interdisciplinary inpatient team conferences and evaluated its effects. Clinician survey impressions were variable, yet favored certain key features introduced by the dashboard. Implementation of the AQ Dashboard also resulted in a large increase in changing the rehabilitation plan, as defined by long-term goal setting in self-care and mobility domains. Patients also met long-term goals in these domains more frequently after implementation of the AQ Dashboard. Post-implementation analysis of LOS and changes in GG scores also indicates improvements in clinical outcomes – improved change scores in a shorter period of time – as a result of use of the AQ Dashboard. Notably, these changes in outcomes surpassed nationally reported change scores and were consistent across different insurance types. Such significant improvements in change scores, coupled with reduced length of stay (a proxy of cost), demonstrates the potential of the AQ Dashboard to enhance value-based care.

Clinician responses favored the AQ Dashboards influence on setting and adjusting goals, tracking patient progress at the domain level, and using predictive models to understand if the patient is on target. These impressions align with several components of precision rehabilitation – specifically that the plan of care for a patient is adjusted based on their progress and prediction to individualize rehabilitation. Likewise, clinicians then adjusted those goals much more frequently and patients attained those goals more often post implementation. Interestingly, clinicians had more negative impressions on the AQ Dashboard’s influence on clinical decisions, facilitating better outcomes for patients, or enhancing goal achievement.

Survey responses also highlighted limitations of the AQ Dashboard regarding its ability to highlight key information for discussion during team conference and identify and address barriers to discharge. The dashboard itself lacks these details - such as, the reason a patient may not be progressing or discharge considerations based on available caregiver support. Such aspects of interdisciplinary team conference must be preserved along with using the AQ Dashboard. Teams also did not agree that it was more efficient to use the AQ Dashboard in team conference.

Determining the specific impacts of the AQ Dashboard on outcomes and LOS is complex and beyond the scope of this project. Recently, Zuang et al. (2022) introduced a framework for evaluating healthcare dashboards, offering clearer guidance on assessing factors influencing interaction effectiveness, user experience, and system efficacy. To ensure the continued effectiveness and scalability of the AQ Dashboard, further evaluation of these factors is necessary to refine its design components.

Teams underwent extensive training to understand and utilize the AQ Dashboard effectively. This training included restructuring team conferences to focus on comparing actual progress with predicted outcomes, identifying specific areas for personalized treatment, and using the predictive model to inform decisions about care plans and length of stay. Scaling this level of training across different institutions would be challenging without dedicated organizational support. Moreover, integrating EHR data into the dashboard requires robust information technology and data science infrastructure.

Another limitation is that our evaluation of the AQ Dashboard was not completed in a controlled environment and did not control for other variables that could affect length of stay, clinical outcomes, or clinician perspective. Future work could evaluate the comparative outcomes of different teams using such a dashboard.

The AQ Metrics address the psychometric flaws of current assessment tools used to document functional progress by accounting for the varying degrees of difficulty and the multidimensionality of assessed functional items. As a continuous scale, it can identify improvement before changes are observable on current assessment tools and provides an avenue for comparison of outcomes between post-acute settings. In contrast to deciphering the many rehabilitation outcome measures that could be used in practice, this composite rehabilitation score is suitable for predictive modeling and relatively easy interpretation by clinical teams.

## Conclusion

We developed, piloted, and measured the impact of a clinical dashboard that incorporated a composite rehabilitation outcome score (cROS), the AbilityQuotient, and group- and patient-specific predictive modeling. Our results demonstrated that such an analytical approach combined with a clinician-friendly dashboard can result in improved patient outcomes and is a step toward precision rehabilitation.

## Data Availability

All data produced in the present study are available upon reasonable request to the authors

## List of Abbreviations

CMS: Center for Medicare and Medicaid
AQ: Ability Quotient
FIM: Functional Independence Measure
IRF: inpatient rehabilitation facility
CFA: confirmatory factory analysis
MIRT: multidimensional item response theory
EHR: electronic health record

## Notes

### Competing Interest Statement

The authors have declared no competing interest.

### Funding Statement

The Shirley Ryan AbilityLab

### Author Declarations

Ethics committee/IRB of Northwestern University waived ethical approval for this work. This work is quality improvement.

