## Supplemental Table 1 for "The AbilityQuotient Dashboard: Outcomes of Implementing Patient-Specific Predictive Modeling in Inpatient Team Conference"

Supplemental Table 1. Self-Care Items

| Assessment Area | Measure | Task | Balance Level  Sitting | Balance Level Standing | Balance Level Walking |
| --- | --- | --- | --- | --- | --- |
| *Balance* | Function in Sitting Test (FIST) | Anterior nudge | **X** | **X** |  |
|  |  | Static sitting | **X** | **X** |  |
|  |  | Sitting with eyes closed | **X** | **X** |  |
|  |  | While sitting, lift foot | **X** | **X** |  |
|  |  | Lateral reach | **X** | **X** |  |
|  |  | Pick up object from floor while seated | **X** | **X** |  |
|  | Berg Balance Scale (BBS) | Standing unsupported |  | **X** | **X** |
|  |  | Standing to sitting |  | **X** | **X** |
|  |  | Standing unsupported with feet together |  | **X** | **X** |
|  |  | Reach forward arm as far as possible while standing |  | **X** | **X** |
|  |  | Pick up object from floor while standing |  | **X** | **X** |
|  |  | Turn around 360 degrees |  | **X** | **X** |
|  | Functional Gait Assessment (FGA) | Gait on level surface |  |  | **X** |
|  |  | Vertical head turns while walking |  |  | **X** |
|  |  | Walk, make pivot turn, and return |  |  | **X** |
|  |  | Step over an obstacle while walking |  |  | **X** |
|  |  | Walking backwards |  |  | **X** |
| Upper Extremity Function (UEF) | Action Research Arm Test (ARAT) | Grasp 10 cm wooden block | **X** | **X** | **X** |
|  |  | Grip 2.25 cm metal tube | **X** | **X** | **X** |
|  |  | Pinch 3 mm ball bearing | **X** | **X** | **X** |
|  |  | Gross movement of hand to mouth | **X** | **X** | **X** |
|  | Bilateral Task Assessment (BTA) | Button 4 buttons | **X** | **X** | **X** |
|  |  | Open toothpaste cap and 4 jar lids | **X** | **X** | **X** |
|  |  | Cut putty with knife into 4 pieces | **X** | **X** | **X** |
|  |  | Fold paper into thirds and place in envelope | **X** | **X** | **X** |
|  |  | Put paperclips on edge of envelope | **X** | **X** | **X** |
|  | 9 Hole Peg Test | Nine hole/peg test, left hand | **X** | **X** | **X** |
|  |  | Nine hole/peg test, right hand | **X** | **X** | **X** |
|  | Box and Blocks Test | Box/Blocks test, left hand | **X** | **X** | **X** |
|  |  | Box/Blocks test, right hand | **X** | **X** | **X** |
|  | Grip Strength | Grip strength, left hand | **X** | **X** | **X** |
|  |  | Grip strength, right hand | **X** | **X** | **X** |
|  | Pinch Strength | Key grip strength with left hand | **X** | **X** | **X** |
|  |  | Key grip strength with right hand | **X** | **X** | **X** |
| Swallowing | Mann Assessment of Swallowing Ability (MASA) | Ability to control salivation | **X** | **X** | **X** |
|  |  | Anterior/posterior tongue mobility | **X** | **X** | **X** |
|  |  | Tongue strength on resistance tasks | **X** | **X** | **X** |
|  |  | Ability to control tongue on repetitive tasks | **X** | **X** | **X** |
|  |  | Ability to break down food/mix with saliva | **X** | **X** | **X** |
|  |  | Ability to swallow chewed food | **X** | **X** | **X** |
|  |  | Time it takes chewed food to be swallowed | **X** | **X** | **X** |
|  |  | Response to cough command | **X** | **X** | **X** |
|  |  | Swallowing function (hypolaryngeal movement) | **X** | **X** | **X** |
|  |  | Vocal quality/presence of coughing post swallow | **X** | **X** | **X** |
|  | Functional Oral Intake Scale (FOIS) | Type of food consistency required by patient | **X** | **X** | **X** |
|  | RIC-DSS | Percentage of time patients need cues to safely manage diet | **X** | **X** | **X** |
| Self-Care Skills | Quality Indicators (QI) | Eating | **X** | **X** | **X** |
|  |  | Oral Hygiene | **X** | **X** | **X** |
|  |  | Shower/Bathe Self | **X** | **X** | **X** |
|  |  | Upper Body Dressing | **X** | **X** | **X** |
|  |  | Lower Body Dressing | **X** | **X** | **X** |
|  |  | Toileting Hygiene | **X** | **X** | **X** |
|  |  | Putting On/Taking Off Footwear | **X** | **X** | **X** |
