## Supplemental Table 2 for "The AbilityQuotient Dashboard: Outcomes of Implementing Patient-Specific Predictive Modeling in Inpatient Team Conference"

Supplemental Table 2. Mobility Items

| Assessment Area | Measure | Task | Mode of Locomotion at DC: Wheelchair | Mode of Locomotion at DC: Walk | Mode of Locomotion at DC: Both |
| --- | --- | --- | --- | --- | --- |
| *Sitting Balance* | Function in Sitting Test (FIST) | Anterior nudge | **X** |  | **X** |
|  |  | Static sitting | **X** |  | **X** |
|  |  | Sitting with eyes closed | **X** |  | **X** |
|  |  | While sitting, lift foot | **X** |  | **X** |
|  |  | Lateral reach | **X** |  | **X** |
|  |  | Pick up object from floor while seated | **X** |  | **X** |
| *Standing Balance* | Berg Balance Scale (BBS) | Standing unsupported |  | **X** | **X** |
|  |  | Standing to sitting |  | **X** | **X** |
|  |  | Standing unsupported with feet together |  | **X** | **X** |
|  |  | Reach forward arm as far as possible while standing |  | **X** | **X** |
|  |  | Pick up object from floor while standing |  | **X** | **X** |
|  |  | Turn around 360 degrees |  | **X** | **X** |
| *Walking* | Functional Gait Assessment (FGA) | Gait on level surface |  | **X** |  |
|  |  | Vertical head turns while walking |  | **X** |  |
|  |  | Walk, make pivot turn, and return |  | **X** |  |
|  |  | Step over an obstacle while walking |  | **X** |  |
|  |  | Walking backwards |  | **X** |  |
|  | Six Minute Walk Test (6MWT) | Six Minute Walk (distance only) |  | **X** | **X** |
|  | Ten Meter Walk Test (10MWT) | Ten Meter Walk (speed only) |  | **X** | **X** |
| *Wheelchair Skills* | Pressure Relief | Pressure Relief | **X** |  | **X** |
|  | Six Minute Push Test | Six minute push (wheelchair) | **X** |  | **X** |
|  | Quality Indicators (QI) | Wheelchair locomotion x 50 ft, 2 turns | **X** |  | **X** |
| *Mobility* | QI | Bed to chair transfer | **X** | **X** | **X** |
|  |  | Walking locomotion x 50 ft |  |  | **X** |
|  |  | Stairs locomotion x 4 steps |  |  | **X** |
|  |  | Walking locomotion x 150 ft |  | **X** |  |
|  |  | Stairs locomotion x 12 steps |  | **X** |  |
|  |  | Toilet transfer | **X** | **X** | **X** |
|  |  | Roll Left/Right | **X** |  |  |
