## Supplemental Table 3 for "The AbilityQuotient Dashboard: Outcomes of Implementing Patient-Specific Predictive Modeling in Inpatient Team Conference"

Supplemental Table 3. Cognitive-Communication Items

| Assessment Area | Assessment | Task | Aphasia | RHD | BI | Cog-comm | Dysarthria/  Voice |
| --- | --- | --- | --- | --- | --- | --- | --- |
| *Memory* | O-log | Name current city |  | **x** | **x** | **x** |  |
|  |  | Name hospital |  | **x** | **x** | **x** |  |
|  |  | Name month |  | **x** | **x** | **x** |  |
|  |  | Name day of the week |  | **x** | **x** | **x** |  |
|  |  | Recall event that resulted in hospitalization |  | **x** | **x** | **x** |  |
|  |  | Recall injury, condition, or deficits |  | **x** | **x** | **x** |  |
|  | 3 Word Delayed Recall | 3 Words |  | **x** | **x** | **x** |  |
|  | RICE-3 | Behavior Observation Profile |  | **x** |  |  |  |
|  | RMBT | Immediate Recall |  |  | **x** | **x** |  |
|  |  | Delayed Recall |  |  | **x** | **x** |  |
| *Pragmatics* | RICE-3 | Pragmatic Communicaion Profile |  | **x** |  |  |  |
| *Agitation* | Agitated Behavior Scale | Frequency exhibits short attention span |  |  | **x** |  |  |
|  |  | Frequency of impulsive behavior |  |  | **x** |  |  |
|  |  | Frequency of uncooperative behavior |  |  | **x** |  |  |
|  |  | Frequency of repetitive behavior |  |  | **x** |  |  |
| *Motor Speech/Voice* | Assessment of Intelligibility of Dysarthric Speech | Intelligible Words |  |  |  | **x** | **x** |
|  |  | Intelligible Words in Sentences |  |  |  | **x** | **x** |
|  | Voice measures | Sustained Phonation Duration |  | **x** | **x** | **x** | **x** |
|  |  | Vocal Intensity during Sustained Phonation |  | **x** | **x** | **x** | **x** |
| *Communication* | BDAE-3 | Basic word discrimination | **x** |  |  |  |  |
|  |  | Follow commands | **x** |  |  |  |  |
|  |  | Complex ideational yes/no | **x** |  |  |  |  |
|  |  | Repeat words | **x** |  |  |  |  |
|  |  | Repeat sentences | **x** |  |  |  |  |
|  |  | Special categories | **x** |  |  |  |  |
|  |  | Match words to pictures (reading) | **x** |  |  |  |  |
|  |  | Oral word reading | **x** |  |  |  |  |
|  |  | Oral sentence reading | **x** |  |  |  |  |
|  | BNT-2 | Total raw score | **x** |  |  |  |  |
| *Writing* | BDAE-3 | Writing legibility | **x** |  |  |  |  |
|  |  | Choice of letters while writing | **x** |  |  |  |  |
|  |  | Ease of writing | **x** |  |  |  |  |
| *Reading* | BDAE-3 | Sentence completion | **x** |  |  |  |  |
|  |  | Paragraph completion | **x** |  |  |  |  |
| *Cognitive-Communication Skills* | Functional Independence Measures (FIM) | Comprehension | **x** | **x** | **x** | **x** | **x** |
|  |  | Expression | **x** | **x** | **x** | **x** | **x** |
|  |  | Social Interaction | **x** | **x** | **x** | **x** | **x** |
|  |  | Memory | **x** | **x** | **x** | **x** | **x** |
|  |  | Problem Solving | **x** | **x** | **x** | **x** | **x** |
